# Differential ventilation using flow control valves as a potential bridge to full ventilatory support during the COVID-19 crisis: From bench to bedside

**DOI:** 10.1101/2020.04.14.20053587

**Authors:** Matthew A. Levin, Martin D. Chen, Anjan Shah, Ronak Shah, George Zhou, Erica Kane, Garrett Burnett, Shams Ranginwala, Jonathan Madek, Christopher Gidicsin, Chang Park, Daniel Katz, Benjamin Salter, Roopa Kohli-Seth, Hung-Mo Lin, James B. Eisenkraft, Suzan Uysal, Michael McCarry, David L. Reich, Andrew B. Leibowitz

## Abstract

**Background:** It has been projected that there will be too few ventilators to meet demand during the COVID-19 (SARS CoV-2) pandemic. Ventilator sharing has been suggested as a crisis standard of care strategy to increase availability of mechanical ventilation. The safety and practicality of shared ventilation in patients is unknown. We designed and evaluated a system whereby one mechanical ventilator can be used to simultaneously ventilate two patients who have different lung compliances using a custom manufactured flow control valve to allow for individual adjustment of tidal volume and airway pressure for each patient.

**Methods:** The system was first evaluated in a simulation lab using two human patient simulators under expected clinical conditions. It was then tested in an observational study of four patients with acute respiratory failure due to COVID-19. Two separately ventilated COVID-19 patients were connected to a single ventilator for one hour. This intervention was repeated in a second pair of patients. Ventilatory parameters (tidal volume, peak airway pressures, compliance) were recorded at five minute intervals during both studys. Arterial blood gases were taken at zero, thirty, and sixty minutes. The primary outcome was maintenance of stable acid-base status and oxygenation during shared ventilation.

**Results:** Two male and two female patients, age range 32-56 yrs, participated. Ideal body weight and driving pressure were markedly different among patients. All patients demonstrated stable physiology and ventilation for the duration of shared ventilation. In one patient tidal volume was increased after 30 minutes to correct a respiratory acidosis.

**Conclusions:** Differential ventilation using a single ventilator and a split breathing circuit with flow control valves is possible. A single ventilator could feasibly be used to safely ventilate two COVID-19 patients simultaneously as a bridge to full ventilatory support.

**Summary Statement:** Not applicable.

## Introduction

The COVID-19 (SARS CoV-2) pandemic has resulted in an unprecedented large number of hospital and intensive care unit (ICU) admissions, with patients requiring mechanical ventilation for prolonged periods of time.^1,2^ Our experience in the Mount Sinai Health System (MSHS) in New York City has been similar, with approximately 10% of all COVID-19 positive patients admitted to date requiring mechanical ventilation, a median duration of ongoing ventilatory support of 9.3 days, 26% mortality despite ventilatory support, and only 25% of patients were extubated successfully (data not shown). This equates to nearly 50% of intubated patients continuing to require ventilatory support. Although the MSHS currently has enough ventilators to meet demand, the slow but steady rise in the number of intubated patients requiring prolonged ventilation is concerning. As this pandemic now spreads throughout the country and around the globe, especially in developing countries, tens of thousands more such patients are expected, leading to even greater concern that there will be insufficient ventilators to meet demand.^3^ The United States Public Health Service Commissioned Corps published a statement that a possible crisis standard of care strategy is the ventilation of two patients with a single mechanical ventilator.^4^ The document described the challenges that must be overcome, and asserted that such a strategy should only be considered as an absolute last resort.

The safety and efficacy of shared ventilation is unknown. A Consensus Statement of six professional organizations including the Society of Critical Care Medicine (SCCM), American Association for Respiratory Care (AARC), American Society of Anesthesiologists (ASA), Anesthesia Patient Safety Foundation (ASPF), American Association of Critical-Care Nurses (AACN), and American College of Chest Physicians (CHEST), advised clinicians that sharing mechanical ventilators “should not be attempted because it cannot be done safely with current equipment” (see Appendix).^5^

Despite these concerns, the Governor of New York, Andrew M. Cuomo, issued a statement at the end of March 2020 approving the use of ventilator sharing as a last resort.^6^ We therefore proceeded to design a novel system and method of shared ventilation that uses a custom manufactured flow control valve to overcome the important challenges of currently proposed systems.

The purpose of this study was to design and build a system that uses a single standard mechanical ventilator to (1) safely ventilate two patients simultaneously, (2) allow individualized setting of tidal volumes and airway pressures, and (3) address the concerns listed in the Consensus Statement by testing it in consented COVID-19 patients as a proof of concept.

## Methods

### Split Circuit

The split circuit system design is as follows (Figure 1). The ventilator breathing circuit is split using standard T-pieces and connectors. Attached to either end of the T-piece is a custom designed and manufactured adjustable flow control valve, followed by a one-way valve, and then the inspiratory limb of the breathing circuit. The flow control valve is an investigational device not yet approved by the Food and Drug Administration. An Emergency Use Authorization has been submitted. At the distal (patient) end, a standard spirometry sensor is placed in-line between the endotracheal tube (ETT) elbow connector and the wye connector of the breathing circuit. The sensor is connected to the gas analyzer-spirometry module of a physiologic monitoring system. At the distal end of each expiratory limb, a one-way valve is placed between the circuit wye connection and the expiratory tubing. The ends of the two expiratory limbs are joined with a T-piece that connects to the expiratory port of the ventilator. Bacterial/viral (B/V) filters are placed between the elbow connector and the ETT, and at the inspiratory and expiratory connection ports on the ventilator.

**Figure 1.**
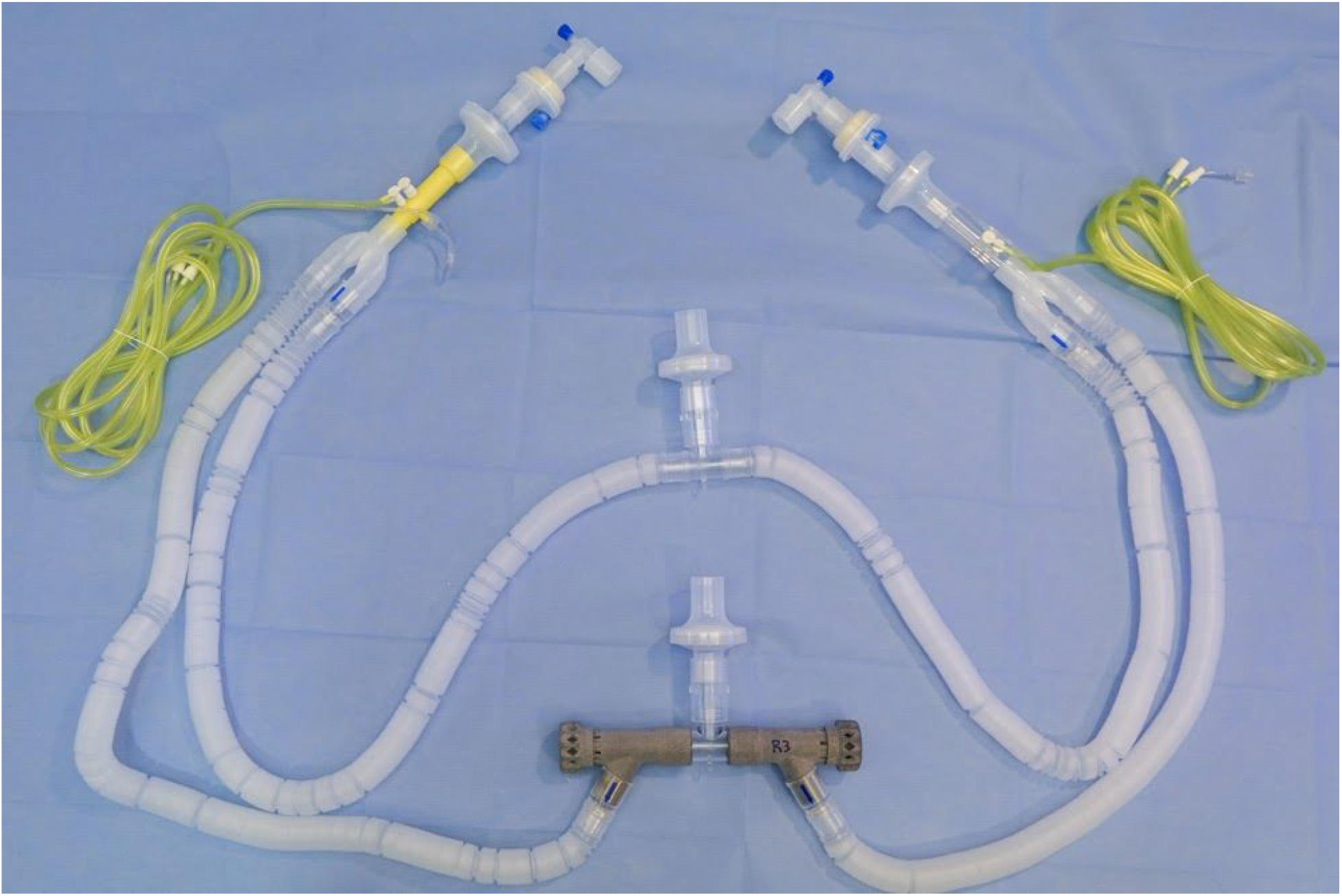
Split Circuit with Flow Control Valves. All components shown are standard breathing circuit components, aside from the flow control valves (in grey) that were 3D printed. The flow control valves are connected to the inspiratory side of the ventilator.

### Simulation Protocol

Two high-fidelity Human Patient Simulator mannequins (HPS Anesthesia Simulator Mannequin Systems, CAE Healthcare, Sarasota, FL) were used for simulation. We tested the system under a variety of simulated patient physiologies likely to be encountered in patients with acute respiratory failure due to COVID-19, using both a GE-Datex-Ohmeda S5 anesthesia machine and a Puritan Bennett 840 ventilator (Medtronic, Minneapolis, MN). The ventilator system checks were run and the split breathing circuit was connected to the two mannequins, designated Mannequin A and Mannequin B. The ventilator was set to Pressure Control mode and both mannequins were set to simulate identical compliance. With both valves open the driving pressure on the ventilator was adjusted to deliver a tidal volume of 4-6 ml/kg ideal body weight (IBW) to Mannequin A.

Mannequin A was then adjusted to the minimum compliance and Mannequin B to the maximum compliance setting available on the simulator. Control valve B was turned in measured increments to a partially closed position until Mannequin B received a similar tidal volume to its baseline. Then, the driving pressure on the ventilator was increased to deliver approximately the same tidal volume to Mannequin A. Thereafter, control valve B was adjusted clockwise to a more closed position to maintain the same tidal volumes for the more compliant Mannequin B. Driving pressure, peak pressure, tidal volume, positive end-expiratory pressure (PEEP), and degree of valve opening were recorded throughout each simulation session experiment.

### Clinical Protocol

The use of this crisis-driven modification of the ventilator breathing circuit was done with the knowledge and agreement of the Program for the Protection of Human Subjects at the Icahn School of Medicine at Mount Sinai. Four patients with acute respiratory failure due to COVID-19 who had already been on mechanical ventilation with sedation and paralysis for 1-12 days, and expected to require continued prolonged ventilatory care, were identified. The patients were required to have similar minute ventilation, PEEP, and fraction of inspired oxygen (FiO_2_) requirements. Informed Consent and Permission for Emergency Procedure with an Unapproved Article was obtained from the legally authorized representative of both patients prior to beginning the study. The patients were moved into the same room in an intensive care unit and connected to full physiologic monitoring. Each patient was placed on pressure control ventilation with the same respiratory rate, PEEP, and FiO_2_. The driving pressure of each patient’s ventilator was adjusted to achieve a tidal volume of 4-6 ml/kg of IBW. The patients were then observed for a period of 20 minutes to ensure they were hemodynamically stable, and a baseline arterial gas sample was taken. Simultaneously, the split ventilation circuit including flow control valves was fully assembled and a third ventilator was set up between the two patient beds and tested with a single circuit. The single circuit was removed and the split circuit was connected.

The shared ventilator was set with the parameters of the patient with the higher driving pressure. Split ventilation was then initiated by clamping the ETT of each patient, disconnecting them from their individual ventilators, and quickly connecting the split circuit. The flow control valve of the patient with the lower driving pressure was quickly throttled down to almost full close, and then slowly opened to achieve similar tidal volumes to the patient’s baseline value. The patients were closely monitored with an intensivist and anesthesiologist at bedside. Thirty minutes after going on the shared circuit, an arterial blood gas was drawn from each patient and the flows to each patient adjusted as needed. After sixty minutes, another arterial blood gas was drawn. At this point, the patients were disconnected from the split circuit by clamping their ETT and reconnected to their individual ventilators. The patients were closely monitored for an additional one hour after termination of the study, in order to ensure that they were stable and had returned to their baseline respiratory status.

## Results

### Simulation Study

The split circuit performed as expected in both simulator scenarios (identical compliance and disparate compliance). Figure 2 shows the flow control valves connected to the inspiratory port of a ventilator in the simulation lab. Results of the simulation are shown in Figure 3. As the flow control valve was progressively opened from a full close position, the title volume delivered to the attached mannequin began to rise in a non-linear but controllable fashion. This was demonstrated across a range of simulated lung and chest wall compliance combinations.

**Figure 2.**
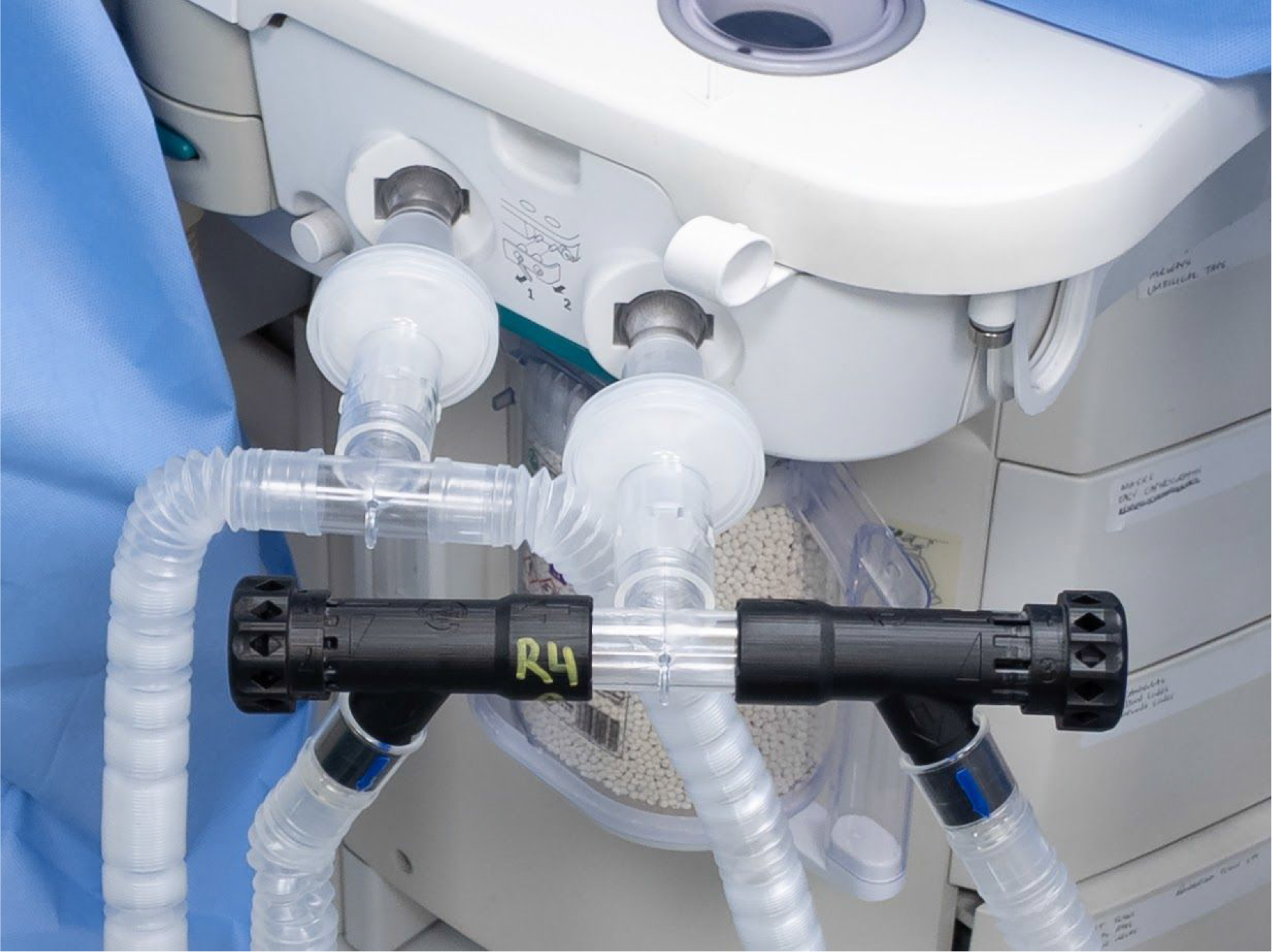
Split Circuit System Connected to a Ventilator. Flow control valves and split circuit connected to a ventilator in the simulation lab, in this case an anesthesia machine ventilator.

**Figure 3.**
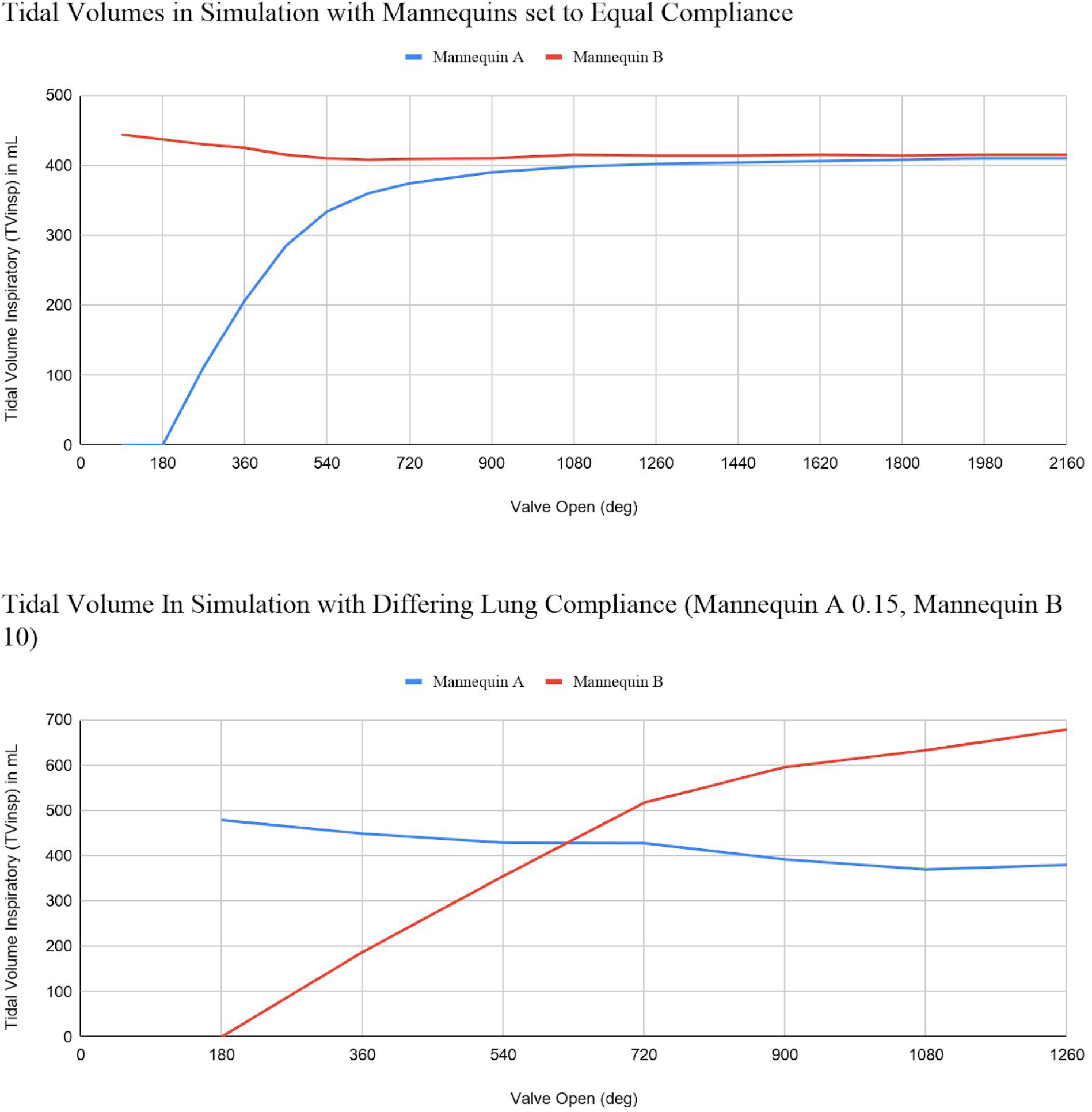
Tidal Volumes During Simulation. Top panel: Mannequins set to equal compliance. Bottom panel: Mannequins with differing compliance. Compliance is a unitless dimension set on the simulator with range 0.15 - 10. Higher numbers indicate greater compliance. The bottom panel demonstrates that tidal volume increased in the more compliant mannequin as the valve was opened, while staying essentially constant in the less compliant mannequin.

### Patient Results

The system performed as predicted by the simulation and there were no adverse sequelae in any of the patients. We were able to successfully ventilate the two pairs of patients for one hour from a single ventilator, with independent adjustment of tidal volume and pressure for each patient. Afterward, each patient was reconnected to a separate ventilator. All study patients remain alive and still ventilator-dependent.

Baseline patient demographics are shown in Table 1. In the first pair, Patient A was a 49-year-old female with an IBW of 46 and Patient B was a 55-year-old male with an IBW of 64. Both patients were sedated and paralyzed for the duration of the study. At baseline both patients were stable on a moderate degree of hemodynamic support (Patient A: norepinephrine 130 ng/kg/min, Patient B: norepinephrine 100 ng/kg/min). The results of an arterial blood gas drawn at time 0 just prior to initiation of split ventilation are shown in Table 2.

**Table 1.**
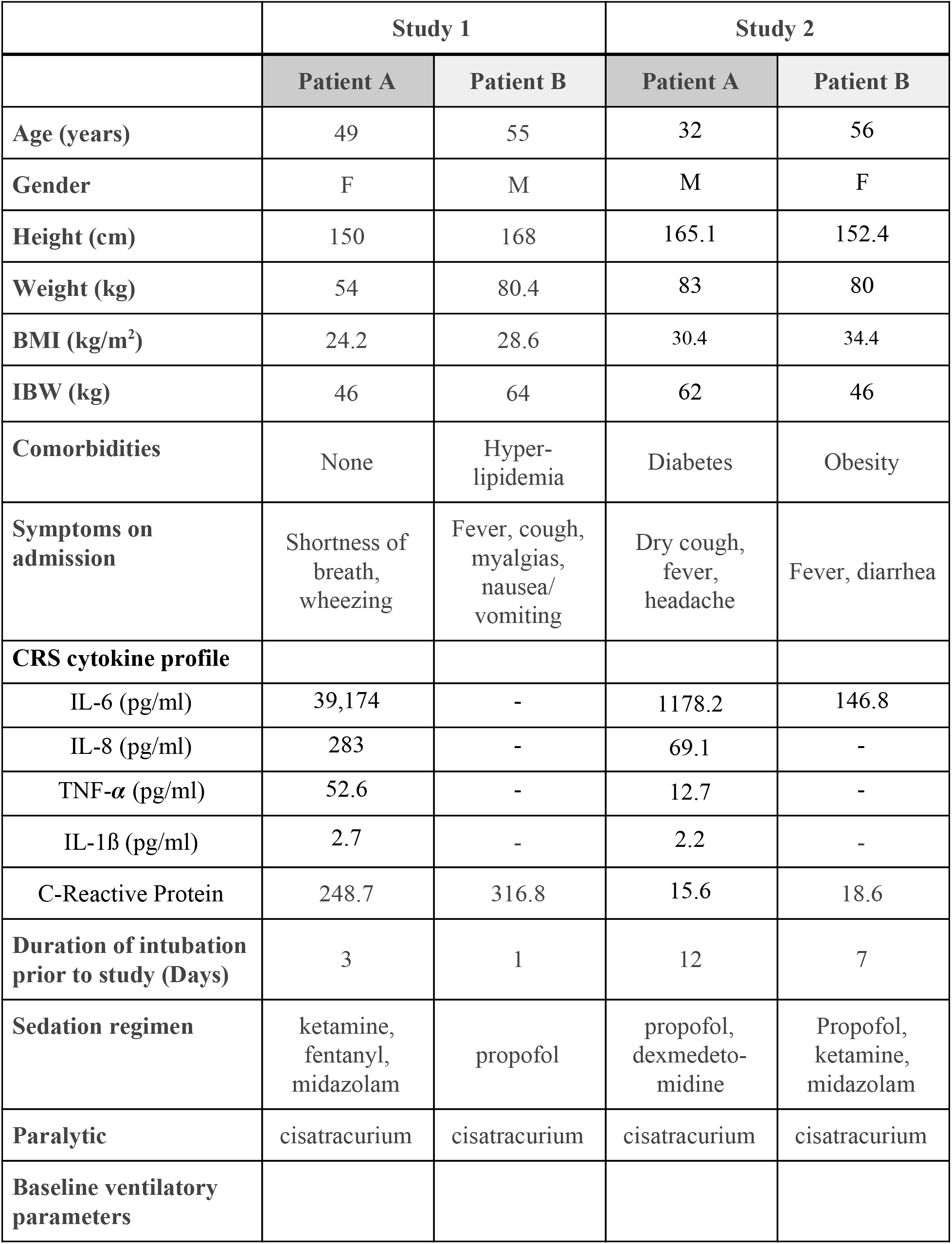
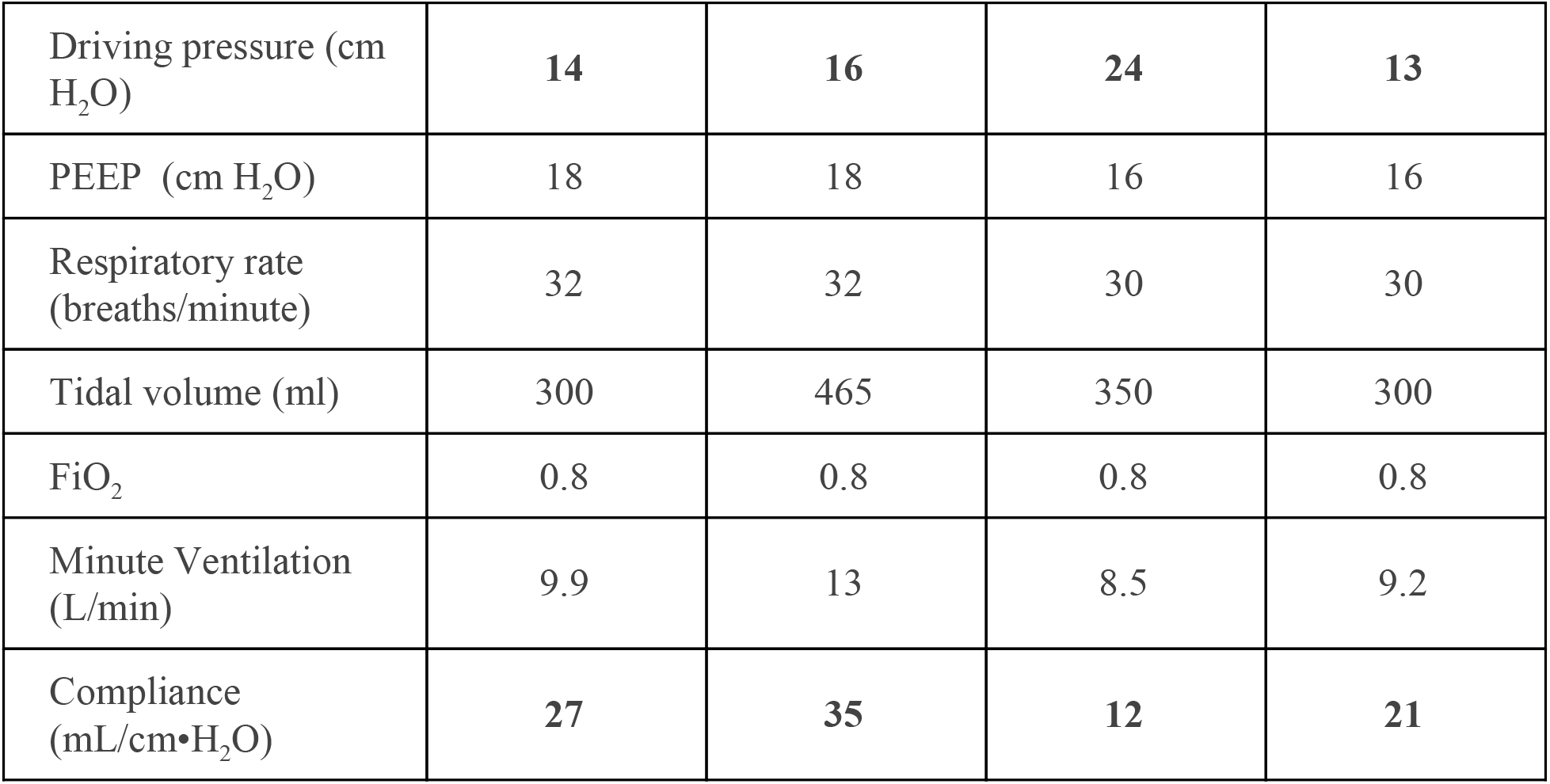
Patient Characteristics.

**Table 2.** Arterial blood gases during the Studies.

|  | Study 1 |  |  |  |  |  | Study 2 |  |  |  |  |  |
| --- | --- | --- | --- | --- | --- | --- | --- | --- | --- | --- | --- | --- |
|  | Patient A |  |  | Patient B |  |  | Patient A |  |  | Patient B |  |  |
| Time (min) | 0 | 30 | 60 | 0 | 30 | 60 | 0 | 30 | 60 | 0 | 30 | 60 |
| pH | 7.37 | 7.25 | 7.35 | 7.33 | 7.34 | 7.35 | 7.38 | 7.32 | 7.33 | 7.42 | 7.33 | 7.36 |
| PaO<sub>2</sub><br>(mmHg) | 131 | 215 | 151 | 154 | 220 | 263 | 83 | 73 | 74 | 138 | 156 | 168 |
| PaCO<sub>2</sub><br>(mmHg) | 52 | 66 | 52 | 47 | 42 | 42 | 57 | 65 | 66 | 44 | 51 | 50 |
| Base<br>Excess<br>(mmol/L) | 4 | 0.7 | 2.4 | -1.6 | -3 | -2.4 | 7 | 5.6 | 6.9 | 3.5 | 0.2 | 1.9 |
| Lactate<br>(mmol/L) | 2.1 | 1.9 | 2.2 | 1.5 | 2 | 2 | 1 | 0.9 | 0.9 | 1.1 | 1.1 | 1.1 |

After baseline arterial blood gas measurements, the patients were switched to the split circuit. The flow control valve on Patient A was slowly dialed down until a tidal volume of ∼6 ml/kg IBW was obtained. Tidal volumes for both patients initially decreased and airway pressures increased (Figure 4). They then remained stable for 30 minutes. An arterial blood gas drawn at 30 minutes demonstrated that Patient A had developed a respiratory acidosis (Table 2). The flow control valve for Patient A was slowly opened until a tidal volume of ∼8 ml/kg IBW was achieved (Figure 4). The patients were observed for another 30 minutes under these new conditions. A final arterial blood gas was drawn, demonstrating that the respiratory acidosis of Patient A had been corrected (Table 2). The patients were then simultaneously reconnected to their original ventilators and the study was terminated.

**Figure 4.**
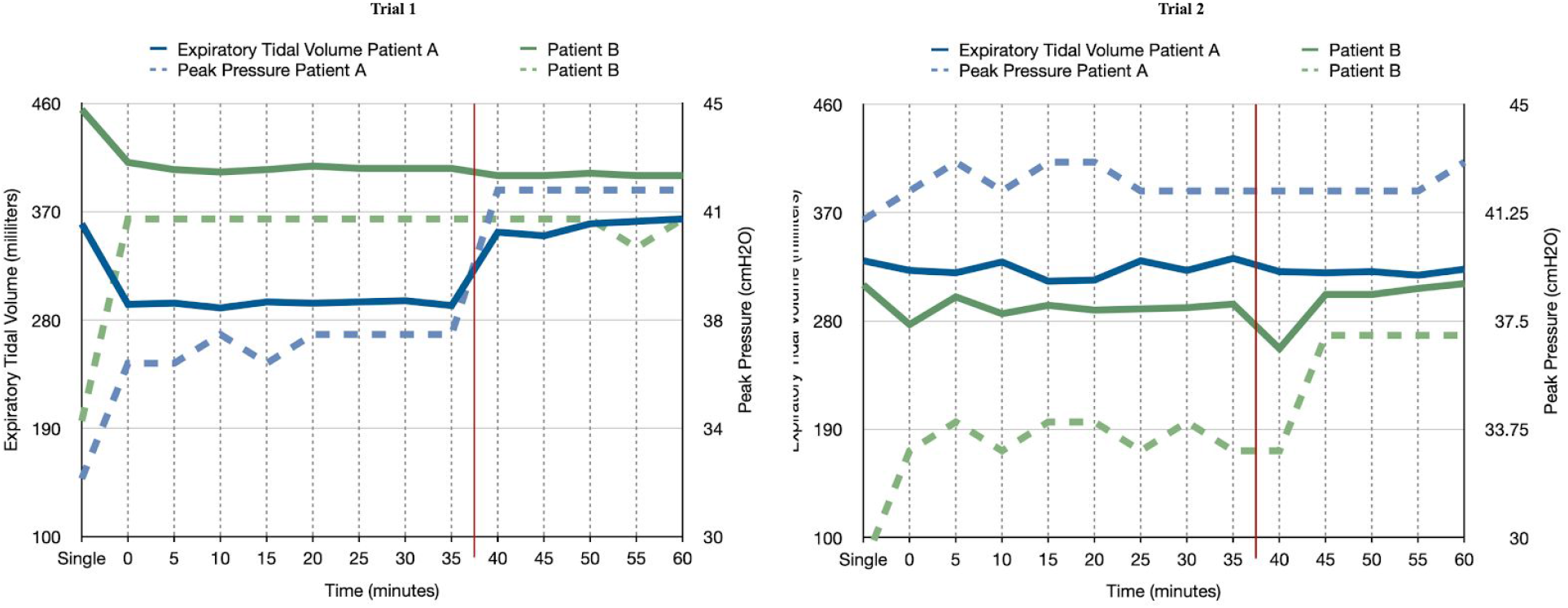
Tidal Volumes and Peak Airway Pressures During the Human studies. Tidal volumes are shown in solid lines and peak airway pressures are shown in dashed lines. Left Panel: Study 1. At the start of the study, tidal volumes fell for both patients, and peak airway pressures rose. Both parameters then stabilized. The red line indicates when the flow control valve for patient A was further opened by ¾ of a turn. The tidal volume and peak airway pressure for patient A subsequently rose, while the tidal volume and peak airway pressure for patient B remained unchanged. Right Panel: Study 2. Tidal volume fell and peak airway pressure rose, as in study 1. The red line indicates valve adjustment for Patient B. In this case, the valve was closed slightly and then opened again to allow slightly more flow. Tidal volume for Patient B remained nearly the same while peak airway pressure rose.

For the second pair, Patient A was a 32-year-old male with an IBW of 62 kg, and Patient B was a 56-year-old female with an IBW of 46 kg. Neither patient was on hemodynamic support. Notably, Patient A had extremely poor lung compliance of 12 mL/cm•H_2_O and required a driving pressure of 24 cm•H_2_O, whereas Patient B was more compliant (21 mL/cm•H_2_O) and required a much lower driving pressure of only 13 m•H_2_O. Once again both patients were ventilated for one hour with no adverse sequelae (Table 2 and Figure 4).

## Discussion

Using a rapid bench-to-bedside approach, we developed a split ventilation circuit that utilizes custom manufactured flow control valves to deliver differential ventilation to two patients connected to a single ventilator. While other groups have proposed similar solutions, we believe that this is the first study to test this technique of using individual flow control valves during shared ventilation in human subjects.^7^ Our study validates that this approach to split ventilation is effective and safe in a small series of COVID-19 patients who differed significantly in size and lung compliance. Both pairs of patients tolerated one hour of shared ventilation with no adverse effects and were alive as of the time of writing. This study demonstrates this technique as a potential bridge to full ventilatory support.

As the number of patients with COVID-19 who require prolonged ventilation increases, the potential lack of mechanical ventilators is worrisome. States and institutions have protocols for allocation of mechanical ventilators such that some salvageable patients by default may receive only palliative care, as exemplified by the New York State Ventilator Allocation Guidelines.^8^ Opportunity for ventilation can be increased if ventilator sharing can be safely utilized. The concept of ventilator sharing is not new, and simulated ventilator sharing has been previously described.^9–12^ However, the only report that describes the use of a valve in a breathing circuit to achieve differential lung ventilation in clinical practice was in a single patient.^13^ At least one recently published protocol provides guidance on shared ventilation, but it has limitations.^6^ Differential ventilation using one ventilator and a split circuit with flow control valves adequately addresses all of the concerns expressed in the consensus statement that recommends against shared ventilation (see Appendix) and has advantages over previous protocols. The adjustable flow valves allow the pressure and volume delivered to each patient to be continuously individualized and titrated to changes in compliance. Appropriately-placed spirometry sensors enable accurate measurement of the delivered tidal volume, airway pressure, compliance, and end-tidal CO_2_ for each patient individually. These data can be displayed on a wall-mounted or portable monitor. The unidirectional valves in both the inspiratory and expiratory limbs prevent reverse gas flow in the circuits or mixing of breathing gas between patients.

### Limitations

Our current design has several limitations. First it does not allow for individualized control of respiratory rate, PEEP, or FiO_2_. In our limited clinical experience, this did not prove to be a significant issue in critically ill COVID-19 patients in the acute phase of their disease. Second, it requires prolonged sedation and paralysis, although many COVID patients require paralysis for optimal ventilation. Third, if one patient in the pair decompensates there may be a significant increase in ventilatory requirement requiring them to be separated emergently from the split circuit and connected to an independent means of ventilation (e.g., Ambu bag). Fourth, even with B/V filters it is possible that bacterial super-infection could be spread between patients. However the U.S. Public Health Service Commissioned Corps guideline on Optimizing Ventilator Use during the COVID-19 Pandemic includes a statement from the Centers for Disease Control (CDC) stating the following with regard to infection control of co-vented patients: “…with the criteria specified and if done with currently established infection control interventions to reduce healthcare-associated infections, including ventilator associated infections, any additional risk is likely to be small and would likely be appropriate in a crisis standard of care.”^14^ Fifth, it will not be possible to wean patients from controlled ventilation while they are on the split circuit. This is a known limitation and we do not expect weaning to be attempted while patients are sharing a ventilator. Finally, our design adds an additional layer of complexity with which not many clinicians will be familiar, thus introducing possible use error that could cause significant harm.

### Conclusion

We have successfully demonstrated that patients of differing size and lung compliance can be independently ventilated with different tidal volumes and pressures using a single ventilator and a split circuit system with flow control valves. The clinical performance of this novel circuit was predicted using high fidelity simulation. We were able to compensate for respiratory acidosis in one patient without affecting the other. All patients remained hemodynamically stable without any negative effect on oxygenation. This approach may be used as a bridge to a next level of therapy, providing acute mechanical ventilation for a short period of time while the next best course of action is determined. Further evaluation is needed to confirm that this system may also be used for longer periods of time. A rapid innovation will be addition of purpose-designed calibrated PEEP valves in the expiratory limbs.

## Data Availability

Data referred to in the manuscript will be made available if asked for.

## Acknowledgments

Not applicable.

## Appendix Response to Consensus Statement on Multiple Patients per Ventilator

Our novel setup addresses the concerns expressed in the Consensus Statement on Multiple Patients per Ventilator.^1^

1. **Volumes would go to the most compliant lung segments**. Placement of control valves in both limbs of the split circuit and measurement of individual patient tidal volumes enables separate control of each tidal volume.
2. **Positive end**-**expiratory pressure, which is of critical importance in these patients, would be impossible to manage**. This can be addressed by choosing patients requiring similar levels of PEEP. If it is determined that PEEP requirements for the patients have begun to diverge significantly, the patients should be removed from split ventilation and placed back on individual ventilators.
3. **Monitoring patients and measuring pulmonary mechanics would be challenging, if not impossible**. Individual patient monitoring is accomplished using separate gas analyzer/spirometry tubing for each patient, connected to separate monitors.
4. **Alarm monitoring and management would not be feasible**. Individual spirometry monitoring alarms are available and can be used.
5. **Individualized management for clinical improvement or deterioration would be impossible**. With a control valve in each inspiratory limb, individualized management is possible-not impossible.
6. **In the case of a cardiac arrest, ventilation to all patients would need to be stopped to allow the change to bag ventilation without aerosolizing the virus and exposing healthcare workers. This circumstance also would alter breath delivery dynamics to the other patients**. Not necessarily. If one patient arrests, the valve on the breathing circuit can be closed, the expiratory limb clamped, and ventilation immediately continued using a self-inflating manual resuscitation device.
7. **The added circuit volume defeats the operational self**-**test (the test fails). The clinician would be required to operate the ventilator without a successful test**,**adding to errors in the measurement**. The ventilator would be tested with a single circuit prior to sharing. Measurements can be individualized.
8. **Additional external monitoring would be required. The ventilator monitors the average pressures and volumes**. Individual monitoring is available.
9. **Even if all patients connected to a single ventilator have the same clinical features at initiation, they could deteriorate and recover at different rates, and distribution of gas to each patient would be unequal and unmonitored. The sickest patient would get the smallest tidal volume and the improving patient would get the largest tidal volume. The greatest risks occur with sudden deterioration of a single patient (e**.**g**., **pneumothorax, kinked endotracheal tube), with the balance of ventilation distributed to the other patients**. The use of flow control valves and individual patient monitoring and alarms overcome these risks.
10. **Finally, there are ethical issues. If the ventilator can be lifesaving for a single individual, using it on more than one patient at a time risks life**-**threatening treatment failure for all of them**. Each institution has their own Crisis Management Policy that should address this.

